# Impact of virus testing on COVID-19 case fatality rate: estimate using a fixed-effects model

**DOI:** 10.1101/2020.04.26.20080531

**Authors:** Anthony Terriau, Julien Albertini, Arthur Poirier, Quentin Le Bastard

## Abstract

**Background:** In response to the SARS-CoV2 pandemic, governments have adopted a variety of public health measures. There are variations in how much testing has been done across countries. South Korea, Germany, and Iceland take the bet of massive testing of their population. Whereas tests were not performed widely in southern European countries. As the former undergo a lower case-fatality rate due to the COVID-19 than the latter, the impact of the testing strategy must be investigated. In this study, we aimed to evaluate the impact of testing on the case fatality rate.

**Methods:** We use data on inpatients across French geographic areas and propose a novel methodology that exploits policy discontinuities at region borders to estimate the effect of COVID-19 tests on the case-fatality rate. In France, testing policies are determined locally. We compare all contiguous department pairs located on the opposite sides of a region border. The heterogeneity in testing rate between department pairs together with the similarities in other dimensions allow us to mimic the existence of treatment and control groups and to identify the impact of testing on mortality.

**Results:** The increase of one percentage point in the test rate is associated with a decrease of 0.001 percentage point in the death rate. In other words, for each additional 1000 tests, one person would have remained alive.

**Conclusion:** Massive population testing could have a significant effect on mortality in different ways. Mass testing may help decision-makers to implement healthcare measures to limit the spread of the disease.

## Introduction

Since it was reported in late December 2019 from Hubei province in China, the severe acute respiratory syndrome coronavirus 2 (SARS-CoV2) has now spread worldwide with more than 2 million confirmed cases by the end of April 2020.^1^ The outbreak reached Europe via Italy at the end of February and quickly affected the entire continent, making Europe the epicenter by mid-March. The World Health Organization (WHO) declared the SARS-CoV2 to be a pandemic in mid-March 2020. While research is still underway to find a curative treatment, the increasing number of severe cases admitted to hospital has raised fears of overburdening the health care systems.

To prevent such a situation, governments have implemented various public health measures such as mobility restrictions, social distancing, or mass screening strategies. On March 16th, the head of the WHO pronounced in favor of massive population tests, because “you cannot fight a fire blindfolded”.^2^ Yet, there is a growing debate about the impact of mass testing on mortality rates.^3^ We have observed strong differences in testing rates between countries; for example, South-Korea, Germany, or Iceland, have undertaken important screening policies and now report low case-fatality rates. On the contrary, countries like Spain or France have restricted access to diagnostic tests for inpatients or health care workers and now report higher mortality rates.^4^ Unfortunately, cross-country comparisons are difficult due to the strong heterogeneity among countries. Even in the United States of America, endowments for medical centers and lockdown strategies are very different from one state to another.

By contrast, France kept a relatively centralized health system but as the epidemic was expanding, the Health Regional Agencies (ARS) were given autonomy in terms of screening strategies implementation; however, at the same time, a strict lockdown approach was instituted for all regions.^5,6^ Among French regions, the main difference in their strategies was the intensity of testing policies. Screening policies and mortality rate might be related to the fact that testing allows authorities to detect and isolate infected people and to prevent them from transmitting the virus; and also enables early treatment, thus increasing the chances of cure.^7^

We propose a novel approach to assess the impact of focused screening strategies on mortality rates, which exploits policy discontinuities at region borders and contiguous department pairs that are located on opposite sides of a region border. This methodology has been used in an economic setting to evaluate the effects of the minimum wage on earnings and employment in the US.^8^

## Materials and methods

### Data sources and procedures

We conducted a retrospective study, with a prospective database, including the total of patients who were admitted to hospital and afterwards discharged, the total of casualties and the total of tests performed for screening COVID-19 infection (RT-PCR) by out-of-hospital medical laboratories. The sample covers the period from 19/03/2020 to 17/04/2020, which corresponds to a lockdown period in France. All the information was provided daily by the French Public Health Agency (Santé Publique France; https://www.data.gouv.fr/). The data was gathered from different geographic areas within France and no other countries were included. We merged this dataset with information on hospital occupancy rates for intensive care units published by the French Ministry of Health (https://www.sae-diffusion.sante.gouv.fr/). Sociodemographic data were extracted from the National Institute of Statistics and Economic Studies (https://statistiques-locales.insee.fr/). Our analysis took place at the department level.

### Fixed-effects model: Empirical strategy

In our study, we exploit the fact that from 14/03/2020, the French government has activated the third stage of the national plan for the prevention and the control of the epidemic, which translates into non-systematic testing of symptomatic individuals. From this date, testing policies were determined at the region level by the Regional Public Health Agencies. We used a fixed-effects model to assess the impact of the number of tests performed over time at a local geographical level (department) on fatality-cases. In fixed-effects models, subjects serve as their own controls, providing a means for controlling omitted-variable bias. Otherwise stated, fixed-effects models allow controlling for time-invariant heterogeneity, i.e. all possible characteristics that do not change over time.^9^

We used two distinct samples: i) The all-department sample (AD sample) that includes 94 departments distributed across 12 regions; ii) The contiguous border department-pair sample (CBDP sample) that contains all the contiguous department pairs that straddle a region boundary. Metropolitan France counts 96 departments. We excluded two departments, Haute-Corse and Corse-du-Sud, that are part of a region, Corsica, that does not share any direct border with others. Among the 94 departments, 69 lie along a region border. As each department may belong to several department-pairs, we have a total of 237 distinct department-pairs. Our strategy consisted in comparing all contiguous department pairs sharing a region border (See Figure 1 for an example) to identify the effect of testing on the case fatality rate. Tests rate and death rates were calculated using the number of RT-PCR tests and the number of deaths related to COVID-19 divided by the number of patients admitted to the hospital, respectively.

**Figure 1:**
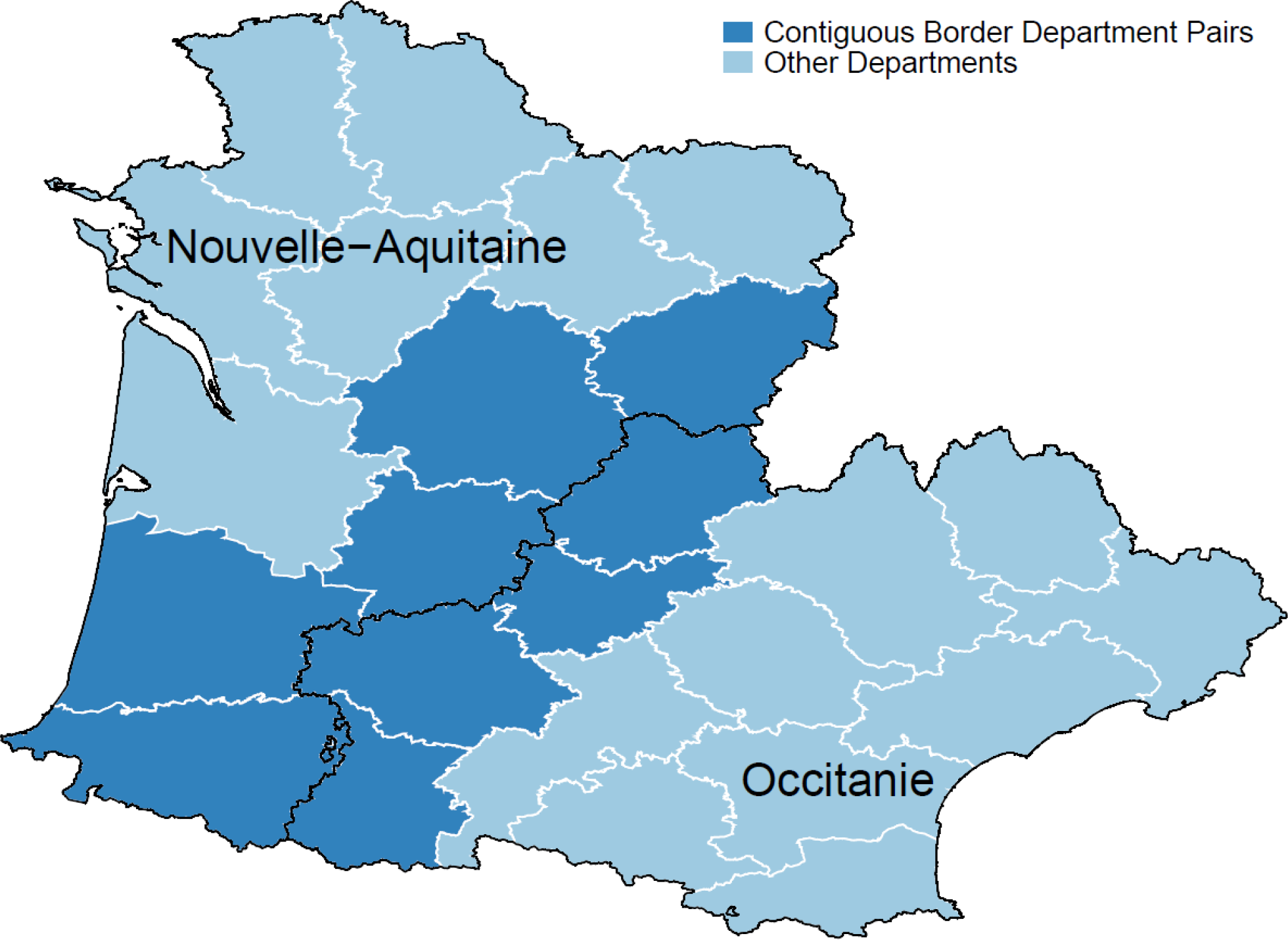
Contiguous border department pairs. Example with “Nouvelle Aquitaine” and “Occitanie” regions.

### Fixed-effects model: Estimation strategy

#### Specifications using the all department sample (AD Sample)

We first estimate the effect of testing on case-fatality rate using the canonical fixed-effects model and the AD sample (Specification (1)):

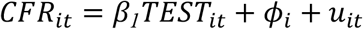

where *i* denote the department, *t* the time, *CFR*_*it*_ is the case fatality rate in department *i* at time *t, TEST*_*it*_ represents the percentage of people hospitalized that are tested in department *i* at time *t, ϕ*_*i*_ is a department fixed effect, and *u*_*it*_ an error term.

#### Specifications using the contiguous border department-pair sample (CBDP Sample)

We now turn to our preferred identification strategy, which exploits policy discontinuities at region borders. To achieve identification, we estimate the following model using the CBDP sample (Specification (4)):

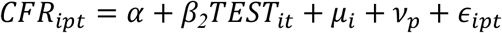

where *i* denote the department, *p* the department-pair, *t* the time, *CFR*_*ipt*_ is the case fatality rate in department *i* in department-pair *p* at date *t, TEST*_*it*_ represents the percentage of people hospitalized that are tested in department *i* in department-pair *p* at date *t, μ*_*i*_ represents a department fixed effect and *v*_*pt*_ a department-pair fixed effect. Standard errors are clustered on the region and border segment separately to account for possible correlation in the residuals.^8^

## Results

### Study Sample

Although fixed-effects models control for all characteristics which do not change over time, we report some time-invariant variables in Table S1 for information. The average number of tested individuals was 613.43 per department with a share of positive cases of approximatively 25 percent. We count a total of 40762 hospitalizations. The observed share of the population above age 65 was roughly 21 percent. As shown by Figure 2, the number of deaths increased quickly, from 154 on March 19^th^ to 11532 on April 17^th^. Over the same period, the number of tests increased from 1713 to 133108.

**Figure 2:**
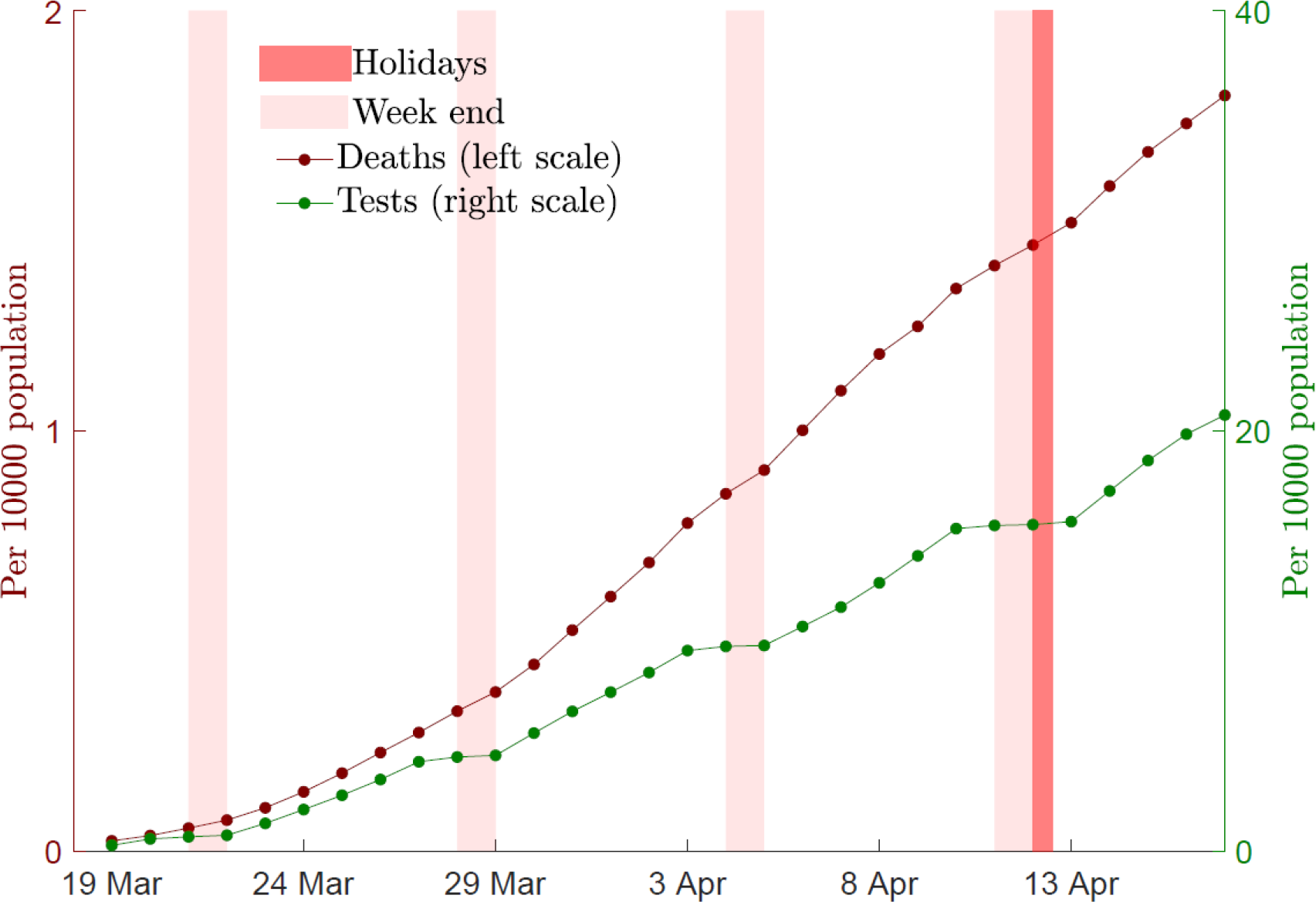
Cumulated number of deaths and tests. Expressed as for 10000 populations. The cumulated number of deaths are those observed in hospitals. The cumulated number of tests are the RT-PCR tests made in private laboratories.

The path of mortality and testing was not homogenous over the territory. The autonomy given to Regional Public Heaths Agencies generated unprecedented differences in testing rate across regions and strong discontinuities at region borders (See Figure 3).

**Figure 3:**
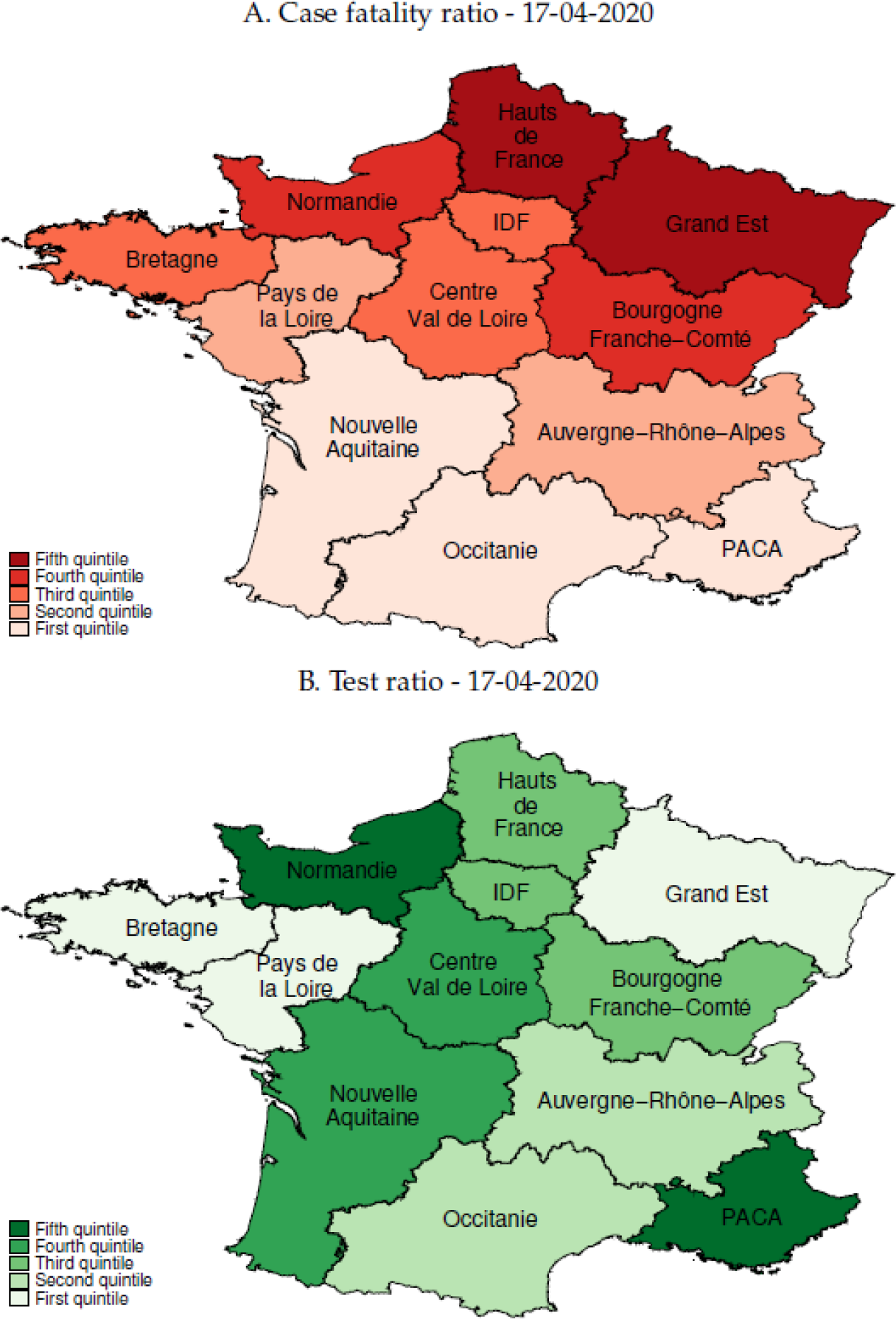
Tests rate and death rates across Region in France. Test rate: number of RT-PCR tests divided by the number of patients admitted to the hospital for COVID-19. Death rate: number of deaths in hospital due to COVID-19 divided by the number of patients admitted to the hospital. We use shapefiles for regions and departement to construct the maps and compute the contiguity matrix (https://www.data.gouv.fr/fr/datasets/contours-des-regions-francaises-sur-openstreetmap/; https://www.data.gouv.fr/fr/datasets/contours-des-departements-francais-issus-d-openstreetmap/). Reading: PACA belongs to the top 20% of regions that test more and to the bottom 20% of regions that have the lowest fatality ratio.

The department fixed effect captures time-invariant heterogeneity across departments. This includes sociodemographic variables (such as the structure of age, race, or gender in the population), but also many variables related to health facilities (number of hospitals, medical density or medical devices). We add time-varying confounding factors in specifications (b) and (c). Specification (b) includes the occupancy rate of the resuscitation units, while specification (c) also controls for the rate of positive tests. The first variable controls for the capacity of hospitals to treat patients at different stages of the COVID-19 epidemic while the second controls for selection bias. Table 1 reports the estimates provided by specifications (a)-(c). Our baseline estimates reveal that a 1 percentage point (pp) increase in the tests/hospitalizations ratio leads to a statistically significant decrease in the case mortality rate by slightly less than 0.001pp.

**Table 1:** Effect of tests on case fatality rate - All-department sample.

| Specification | (a) | (b) | (c) |
| --- | --- | --- | --- |
| Test rate | -0.000849***<br>(0.000281) | -0.000878***<br>(0.000279) | -0.001009***<br>(0.000286) |
| **Controls** |  |  |  |
| Occupancy rate (intensive care unit) | NO | YES | YES |
| Rate of positive tests | NO | NO | YES |
| Number of periods | 30 | 30 | 30 |
| Number of departments | 94 | 94 | 94 |
| Number of regions | 12 | 12 | 12 |

Standard errors between parentheses. \*\*\* p<0.01, \*\* p<0.05, \* p<0.1. Source: Santé Publique France and authors calculations.

Finally, table 2 displays the results for specification (d)-(f). Our estimates reveal that a 1 pp increase in the tests/hospitalizations ratio leads to a statistically significant drop of case mortality rate by 0.001 pp. Putting these numbers into perspective involves that for each additional 1000 tests, one person would have remained alive.

**Table 2:** Effect of tests on case fatality rate - Contiguous border department-pair sample.

| Specification | (d) | (e) | (f) |
| --- | --- | --- | --- |
| Test rate | -0.001043**<br>(0.000516) | -0.001069**<br>(0.000508) | -0.001204**<br>(0.000533) |
| **Controls** |  |  |  |
| Occupancy rate (intensive care unit) | NO | YES | YES |
| Rate of positive tests | NO | NO | YES |
| Department-pair × dummies | YES | YES | YES |
| Number of periods | 30 | 30 | 30 |
| Number of departments | 69 | 69 | 69 |
| Number of department-pairs | 237 | 237 | 237 |
| Number of regions | 12 | 12 | 12 |

Standard errors between parentheses. \*\*\* p<0.01, \*\* p<0.05, \* p<0.1. Source: Santé Publique France and authors calculations.

## Discussion

SARS-CoV-2 outbreak is one of the major public health emergencies of international concern for decades. Countries have implemented various measures mostly based on mobility restriction, social distancing, and regional or national lockdown. All of these public health measures are aimed at “flattening the curve” of the infected cases to limit avoidable mortality due to overburdened health care systems. We evaluate the effect of mass screening COVID-19 on mortality rate in France during the first month of the lockdown. We take advantage of the difference in screening intensity among French regions. We first estimate the effect of testing on case-fatality rate using the canonical fixed-effects model and the AD sample and find that the increase of screening rate of 1 pp allows mortality rate to decrease of nearly 0.001 pp. We confirmed our results by estimating the fixed-effects model using the CBDP sample which compares contiguous French departments sharing region borders.

To the best of our knowledge, no large randomized controlled trial (RCT) has been implemented to investigate the effect of tests on the case-fatality rate, probably due to time, budget, or ethical constraints. When RCT are difficult to implement or unethical, natural experiments (NE) are one of the best alternatives. The principle of NE is to mimic the existence of treatment and control groups using an instrumental variable that induces a change in the explanatory variable but has any direct effect on the outcome. However, in the case of the COVID-19 epidemics, finding a suitable instrument remains a hard task. In the absence of RCT or NE, many researchers try to approximate using standard methods such as linear regression, logistic regression, or propensity scores. However, such methods are subject to the well-known omitted-variable bias, leading to severe bias in estimating the effects of the variables that are included. Consequently, causal inference via statistical adjustment represents a poor alternative to randomized experiments. In such a context, panel data models represent the best way to control for heterogeneity and to improve causal estimation.^8^ We use a fixed-effects model because it represents a powerful tool for longitudinal data analysis.^9^ However, such a model requires substantial differences between treatment intensities across entities and time to get precise estimates. Our data meets these conditions: i) no region has the same test rate path than other regions over the period considered; ii) the test rate varies greatly across regions and time. Methods based on regional controls and policy discontinuities have several advantages: i) contiguous border departments are relatively similar, in particular with regard to health trends, which are of major importance in the context of an epidemic; ii) the testing policy is determined at the region level and is largely exogenous from the point of view of a department, which rules out potential reverse causality.^8^

Until a vaccine is developed, the only way to prevent an unrestrained scenario is to control the spread of SARS-CoV-2. This is a challenging task because some asymptomatic infected patients could potentially spread the virus. Literature reports an alarming proportion of asymptomatic infected cases. Epidemiological data from the Diamond Princess cruise sheep revealed only 18% of positive cases reported no symptoms.^10^ Two hospitals in New York implemented universal testing for SARS-CoV-2 with nasopharyngeal swabs in women who were admitted for delivery, and revealed that nearly 90% of patients who were positive for SARS-CoV-2 at admission reported no symptoms.^11^ Overall population screening in Iceland revealed that only 57% of participants with positive tests reported symptoms of Covid-19.^12^ This proportion could be even higher, because of false negative results of tests to detect SARS-CoV-2.^13^ Testing is part of a strategy to limit the transmission of the virus and WHO recommends a rapid diagnosis and isolation of cases in combination with a rigorous tracking and precautionary self-isolation of close contacts. Several authors support the implementation of mass screening policies.^3,14^ In our opinion, mass screening may positively impact the fatality case rate in different ways. First, unfocused testing, i.e. not limited to symptomatic subjects, could improve the monitoring of the progress of the epidemic and facilitate decision-making by the health authorities. The use of “case definition”, given the limited knowledge of the new disease, probably resulted in a low sensitivity to detect infected subjects, resulting in a delayed perception of the progression of the epidemic.^15,16^ Screening strategies are subject to the availability of tests which indirectly shapes epidemic curves.^17^ While the USA increased their screening capacities between late-February to early-March, the country experienced a rapid increase of total infected cases.^18^ Second, mass screening may also allow early identification of infected subjects and rapid implementation of isolation measures. Early reports from Wuhan suggest that public health interventions combining universal symptoms survey, traffic restriction and home quarantine resulted were temporarily associated with an increased control of the outbreak.^19,20^ A modeling from Singapore suggests that quarantining of infected individuals and their family members, school closure and workplace distancing could reduce the progression of the epidemic but is associated to a significant economic cost.^21^ Review from the Cochrane database concludes that quarantine is important in reducing the number of covid-19 cases but is dependent on screening strategies.^22^ Also, a US survey on the impact of school closure on mortality reports that the transmission prevention by school closure needs to be weighted with the potential loss of health-care workers.^23^ This supports that public health decisions should be as focused as possible in order to limit the negative impact on the economy and the society.^24^ Importance of rapid diagnosis and case identification and isolation will become of utmost importance with the end of lockdowns.

Our study far supports a significant impact of screening strategies on the case-fatality rate in France. Notwithstanding, there are some limitations to our results. First, they belong to France and it would be very hazardous to pretend that they apply to other countries because their exposition to COVID-19 is different, they adopted different strategies, and have different health structures. Second, to provide further evidence on this relation, it would be worth applying this methodology to other countries for which such data are available and in which testing policies are sufficiently heterogeneous across geographical areas.

In addition, the data on tests collected by the French Public Health Agency are those made by private laboratories and do not include those made in public hospitals. This represents an important share of tests (between half and two thirds) and we cannot rule out the possibility that this unobservable amount of screening activity may affect our results. Lastly, our study cannot quantify the respective contribution of the treatment delivered to screened and infected individuals or the lower dissemination of the virus that results from quarantining policies.

## Conclusion

COVID-19 intensive screening policies were significantly associated with a decrease in the fatality-case rate in France. These results support the implementation of mass screening strategies and could provide important information for decision-makers in the fight against SARS-CoV2 pandemic. The optimal testing strategy might also concern economic issues. Indeed, the Bank of France estimated that each fortnight of lockdown costs to France 1.5% of annual GDP (nearly USD48 billions).^25,26^ From a costs/benefits perspective one might naturally wonder what is the optimal policy capable of containing the outbreak and lowering the fatality rate. This is in our research agenda.

## Data Availability

All data used in this manuscript are publicly available online.

https://www.data.gouv.fr/

https://www.sae-diffusion.sante.gouv.fr/

https://statistiques-locales.insee.fr/

## Aknowledge

We thank Dr Natalia Lucia Gomez, from the Hospital Italiano de Buenos Aires, for it’s attentive revision of the manuscript and Pr Emmanuel Montassier, from the University of Nantes for his expert advices.

---

**Table S1:** Descriptive statistics.

|  | All-department<br>Sample |  | Contiguous border<br>departement-pair<br>sample |  |
| --- | --- | --- | --- | --- |
|  | Mean | s.d. | Mean | s.d. |
| *Variables related to individual health* |  |  |  |  |
| Number of people hospitalized | 433.64 | 785.26 | 405.32 | 728.12 |
| Number of tests | 613.43 | 1293.99 | 572.43 | 1141.69 |
| Number of positive tests | 155.71 | 361.35 | 152.45 | 345.93 |
| Number of patients cured | 154.55 | 285.52 | 145.20 | 265.71 |
| Number of deaths | 53.71 | 113.22 | 49.58 | 102.51 |
| *Variables related to health facilities* |  |  |  |  |
| Number of hospital beds (intensive care unit) | 52.30 | 63.33 | 47.30 | 55.92 |
| Number of hospital days (intensive care unit) | 16704 | 21270 | 15124 | 18655 |
| Occupancy rate (intensive care unit) | 107.84 | 43.67 | 109.21 | 45.73 |
| *Sociodemographic variables* |  |  |  |  |
| Surface area (in km<sup>2</sup>) | 52057 | 22718 | 52183 | 22994 |
| Population density | 574.86 | 2453.53 | 500.79 | 2106.07 |
| % of people aged 65 and over | 21.16 | 3.78 | 21.18 | 3.93 |
| Household size | 2.18 | 0.12 | 2.19 | 0.13 |
| Poverty rate | 14.54 | 3.02 | 14.44 | 2.89 |
| Number of periods | 30 | 30 | 30 | 30 |
| Number of departements | 94 | 94 | 69 | 69 |
| Number of department-pairs | NA | NA | 237 | 237 |
| Number of regions | 12 | 12 | 12 | 12 |

Note: Sample means are reported for all departments in France and for all contiguous border department-pairs with a full balanced panel of observations. Source: Santé Publique France

